# Significance testing for small annotations in stratified LD-Score regression

**DOI:** 10.1101/2021.03.13.21249938

**Authors:** Katherine C. Tashman, Ran Cui, Luke J. O’Connor, Benjamin M. Neale, Hilary K. Finucane

## Abstract

S-LDSC is a widely used heritability enrichment method that has helped gain biological insights into numerous complex traits. It has primarily been used to analyze large annotations that contain approximately 0.5% of SNPs or more. Here, we show in simulation that, when applied to small annotations, the block jackknife-based significance testing used in S-LDSC does not always control type 1 error. We show that the inflation of type 1 error for small annotations is due both to the noisiness of the jackknife estimate of the standard error and to the non-normality of the regression coefficient estimates. We use the percent of 0.01 centimorgan blocks in the genome overlapped by the annotation to quantify the size of an annotation and the extent to which the SNPs in the annotation cluster together, and we find thresholds on this value above which type 1 error is controlled. We have implemented a test in the LDSC software that informs users when they compute LD scores for an annotation if the annotation does not pass the threshold for producing controlled type 1 error.

**Author Summary:** Genetics is a rapidly evolving field that allows us to link our genetic code to the physiological manifestations of disease. A key part of this work is finding regions of the genome that contribute disproportionately to the genetic underpinnings of a disease. A commonly used tool to provide such insight is stratified LD score regression (S-LDSC). S-LDSC allows us to estimate how much a set of genomic regions contributes to the overall heritability of a phenotype, and to test whether this is more than we would expect by chance. Here we show that when we apply S-LDSC to a small set of genomic regions, it does not give an accurate test of whether this set of genomic regions contributes more than we would expect by chance to the phenotype. We characterize what it means to be a “small” set of genomic regions, and we set thresholds to restrict which annotations we test to prevent false positive results.This helps to ensure that as we continue to pursue genetic analyses at scale, we report only truly significant results that will help us further understand the etiology of many of the traits we study.

## Introduction

Genome-wide association studies of complex traits have yielded thousands of associated variants; however, such results rarely point to conclusive biological mechanisms. A commonly used tool to gain insight into the biological underpinnings of these traits is enrichment analysis. Stratified LD score regression (S-LDSC)^5^ is a widely used enrichment method that estimates the heritability enrichment of a functional annotation, defined as the proportion of heritability explained by the annotation divided by the proportion of SNPs in the annotation. S-LDSC also estimates the contribution of an annotation to per-SNP heritability in a joint model with other annotations. S-LDSC uses the block jackknife^6^, a commonly used statistical tool in population genetics^1,2,3,4^, for standard error estimation and significance testing.

S-LDSC is typically applied to large annotations covering approximately 0.5% of SNPs or more. Here, we show in simulation that when S-LDSC is applied to small annotations, block jackknife-based significance testing does not always control type 1 error, especially for less polygenic traits. Specifically, we use the percent of 0.01 centimorgan blocks in the genome overlapped by the annotation as a way to quantify the size of an annotation and the extent to which the SNPs in the annotation cluster together, and we find thresholds on this value above which type 1 error is controlled and below which there can be inflation of type 1 error for the different statistical tests conducted by S-LDSC. We then show that for annotations that do not pass this threshold, the type 1 error inflation can be explained by a combination of non-normality of the test statistic and noisiness of the jackknife estimate of the standard error. We have implemented a test in the LDSC software that informs users when they compute LD scores for an annotation if the annotation does not pass the threshold for producing controlled type 1 error.

## Methods

We implemented a simulation framework using 50,000 white British individuals from the UK Biobank and 9.3 million imputed variants to produce three sets of 1250 phenotypes with 200, 1000, and 10000 causal SNPs, respectively. We simulated heritable phenotypes, with h^2^ = 0.6. The causal SNPs and their effect sizes were chosen independently of any annotation, so the true value of *τ*_*c*_ was 0 for every annotation *c* except the base annotation containing all SNPs, and every simulated phenotype.

We created 94 annotations with varying size, varying number of jackknife blocks overlapped out of 200 total, and varying average segment length. To create the annotations, we randomly sampled either a set of genes or a percentage of SNPs from the genome from a specified number of jackknife blocks. For example, if we were to create an annotation of 100 genes that overlapped 30 blocks, we would randomly sample 100 genes from 30 of the default 200 jackknife-blocks such that at least 1 gene came from each block. We simulated annotations with 0.25, 0.1, 0.5, 1, 2, 3, or 5 % of SNPs, or 10, 30, 100, 200, 300, 450, or 600 genes, contained in 2, 4, 6, 8, 10, 30, 60, 80, 100, 150 or 200 jackknife blocks, excluding impossible combinations of parameters such as 5% of SNPs in 2 jackknife blocks. In addition to these simulated annotations, we included 100 real gene sets sampled from MSigDB as well as the 52 annotations in the baseline_v1.1 model^5^, leading to a total of 246 annotations (Table S2).

For each simulated phenotype and each annotation, we ran S-LDSC with the annotation plus the baseline_v1.1 model and calculated both one-sided and two-sided p-values for 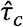 as well as two sided p-values for the heritability enrichment estimates. For each of these three tests, we assessed type 1 error control, in aggregate over annotations and simulated phenotypes, in three ways: first, using the proportion of rejections at P=0.05; second, defining a false positive as anything that passed significance after Bonferroni correcting for the 3750*246 hypotheses tested; and third, by visual inspection of a Q-Q plot.

## Results

S-LDSC is a method for modeling the contributions of functional annotations to heritability using summary statistics. The method jointly models tens of overlapping annotations and reports two types of output: first, a joint-fit regression coefficient estimate for each annotation c, denoted 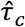, that quantifies the contribution of that annotation to per-SNP heritability; and second, the total heritability explained by SNPs in the annotation, including heritability attributable to other overlapping annotations. The former is typically used to identify phenotype-relevant tissues and cell types, while the latter is used to estimate heritability enrichment. S-LDSC uses the block jackknife to estimate standard errors and error covariance for the vector of estimates 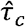 of *τ*_*c*_ The standard errors are then used to compute z-scores to test either the null hypothesis that*τ*_*c*_ ≤0 (one-sided test) or the null hypothesis that *τ*_*c*_ =0 (two-sided test). To test the null hypothesis of no heritability enrichment -- i.e., to test the null hypothesis that the proportion of heritability in a category equals the proportion of SNPs in that category -- the null hypothesis is transformed into a linear condition on *τ* and the jackknife covariance of 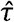 is used to test the null hypothesis.

We used S-LDSC to test the null hypothesis of no heritability enrichment, the null hypothesis that *τ*_*c*_≤0 (one-sided test), and the null hypothesis that *τ*_*c*_ =0 (two-sided test) for each of 3,750 phenotypes simulated with no functional enrichment (true *τ*_*c*_ =0) and each of a set of 246 annotations including many annotations much smaller than is typical for S-LDSC input (see Methods).

Aggregated over all 3,750 simulated phenotypes and over all 246 annotations, our analyses showed controlled type 1 error at a cutoff of P=0.05 but an inflation of very small P-values. Specifically, 3.14% of results were significant at P=0.05, but 78 results passed Bonferroni correction for all annotations and all phenotypes (P < 0.05/(3750*246)) and so were called false positives. Q-Q plots showed extreme inflation of type 1 error for both the one-sided test of *τ*_*c*_ ≤ 0 and the two-sided test of *τ*_*c*_ =0, and very mild inflation of type 1 error for the two-sided test of heritability enrichment =1 (Figure 1). While results were mostly consistent across polygenicities (Figure S1), we observed more false positives for the two least polygenic settings than for the more polygenic setting for the two-sided test (Figure S2). Restricting to the annotations of the baseline model, we found controlled type 1 error for all three tests (Figure S3).

**Figure 1:**
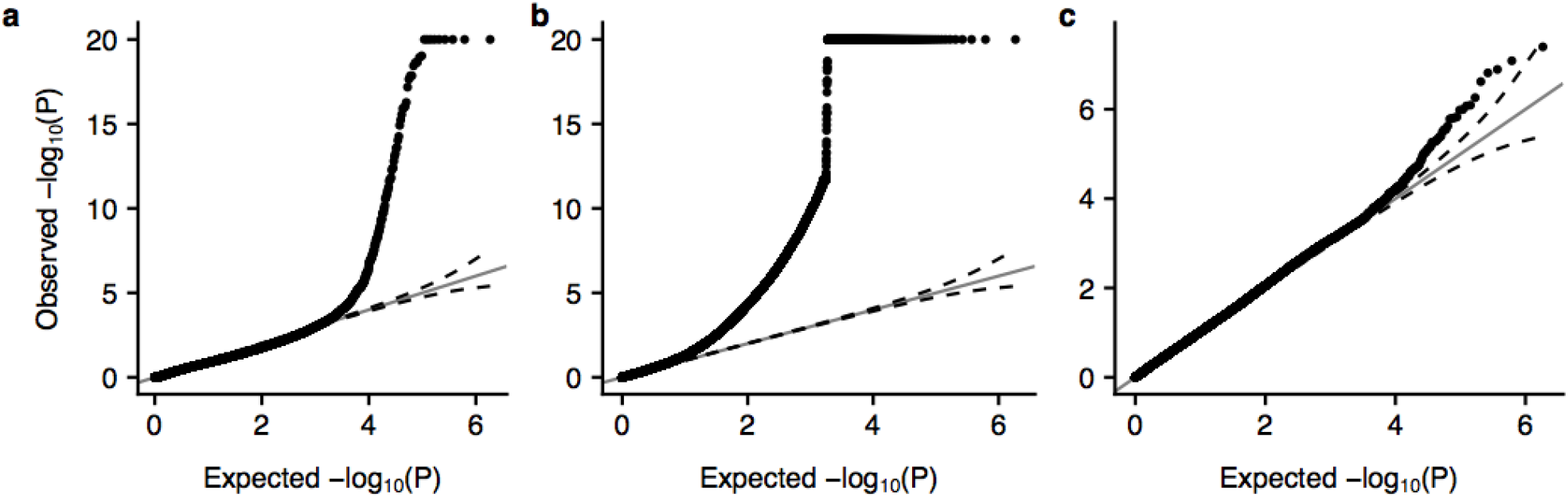
Results of null simulations, aggregated over annotations and simulated phenotypes. Q-Q plots for the **(a)** one-sided test of tauhat <= 0, **(b)** two-sided test of tauhat = 0, and **(c)** two-sided test of heritability enrichment.

We anticipated that type 1 error control for an annotation would depend on both the number of variants in an annotation and the extent to which they cluster together. Thus, we characterized each annotation using six metrics: (1) the percent of SNPs in the annotation; (2) the percent of 0.01 centimorgan (cM) blocks of the genome overlapped by the annotation; (3) the percent of cM blocks of the genome overlapped by the annotation; (4) the percent of 1 cM blocks of the genome overlapped; (5) the percent of jackknife blocks (average size = 18 cM) overlapped by the annotation; and (6) the effective number of independent SNPs10 in the annotation, defined as 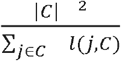 where *l* (*j,C*) is the LD score of SNP *j* to annotation *C*.

For each characterization metric except for number of jackknife blocks overlapped, we found that the most significant P-values tended to occur for annotations with smaller values of the metric: annotations with false positives, on average, had fewer SNPs, overlapped fewer 0.01, 0.1, and 1 cM blocks, and had a smaller effective number of independent SNPs. Specifically, for each annotation, we computed the minimum P-value obtained with the annotation for any of the simulated phenotypes, and for each characterization metric we plotted these minimum P-values across all annotations against the value of the metric (Figure 2a, S4). We then computed the mean value of each metric within annotations with false positives and among all annotations, and found that the mean value of the metric was smaller for annotations with false positives than for all annotations for each of the three statistical tests and five of the six metrics considered (all except for number of jackknife blocks overlapped). Overall, the two-sided test of *τ*_*c*_ =0 had worse type 1 error inflation than the other two tests, and the inflation was captured less well by the six metrics than the other two tests.

**Figure 2:**
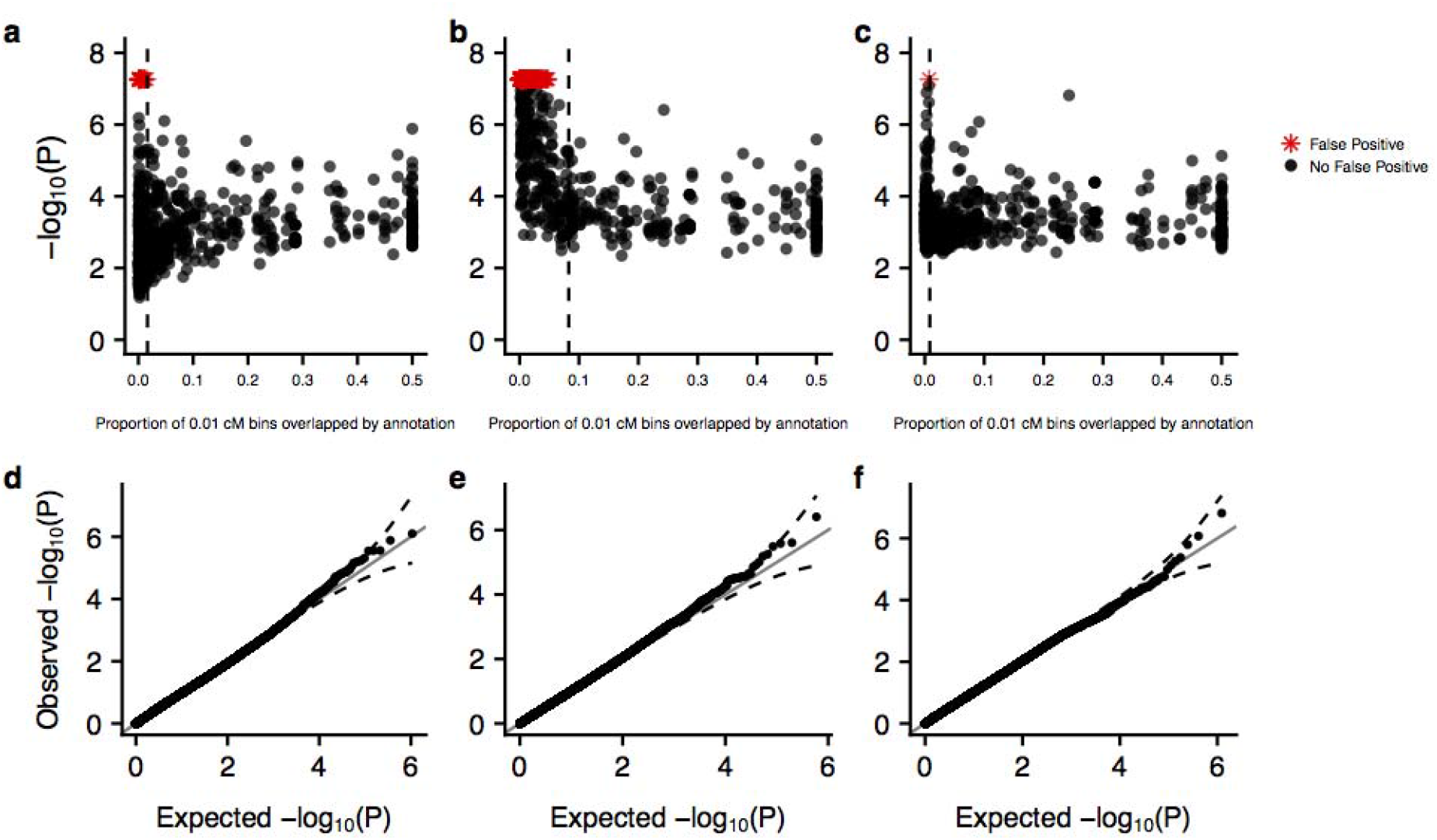
Control of type 1 error is restored by restricting to annotations overlapping sufficiently many 0.01 cM bins. (**a**,**b**,**c**) Dependence of type 1 error on the proportion of 0.01 cM bins overlapped by the annotation for the one-sided, two-sided, and enrichment test respectively. Each dot represents one of the 246 annotations tested. Any p-value below the false positive threshold of P < 0.05/(3750*246) was set to that value for visualization and denoted with a red star. The dashed black line indicates the threshold used to recover a well controlled type 1 error. All annotations overlapping more than 50% of 0.01 cM bins were thresholded to 50% for visualization. (**d**,**e**,**f**) Q-Q plots for the one-sided, two-sided, and enrichment test respectively, restricting to annotations that pass the threshold depicted in **a, b**, and **c**, respectively.

We then evaluated each metric as a diagnostic tool that could be used to exclude annotations with inflated type 1 error. For each characterization metric and each of the three tests, we found the threshold such that excluding all annotations with values of the metric below the threshold would exclude all false positives. We then counted the number of remaining annotations; more remaining annotations indicates that the characterization metric would make a more specific diagnostic tool (Figure S5). We found that results differed for different characterization metrics and for the three tests, but that the percent of 0.01 cM blocks overlapped was a good metric for all three tests (number of remaining annotations = 132, 93, and 161 for the one-sided, two-sided, and enrichment test at a threshold of 1.7, 4.9 and 0.83 percent of 0.01 centimorgan blocks overlapped, respectively.) For the one-sided test and the heritability enrichment test, these thresholds restored type 1 error control. For the two-sided test, a more stringent threshold of 8.3% was needed to restore type 1 error control. We note that the annotations of the baseline model all pass the thresholds for the one-sided and enrichment tests, which are the two most commonly used tests. Because we have shown that type 1 error is controlled for all three tests when restricting to the baseline annotations (Figure S3) we do not recommend excluding annotations from the baseline model even when performing the two-sided test.

Having found that excluding annotations that overlap a small percentage of 0.01 cM blocks of the genome restores type 1 error control, we next sought to understand the source of type 1 error inflation for the annotations that do not pass this threshold. To do this, we focused only on the one- and two-sided test for 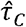,and we considered the z-score of the coefficient used to test for significance in these cases. The z-score of the coefficient is equal to the estimate of the coefficient divided by the estimated standard error. This z-score will have the correct null distribution if the estimate of the coefficient is unbiased and normally distributed, and if the estimated standard error is equal to the true standard deviation of the estimate. We found that the coefficient estimate and standard error estimate were both approximately unbiased (Figure S6), and so we focused on noisiness of the standard error estimate and non-normality of the coefficient estimate as potential explanations for the type 1 error inflation.

To investigate the effect of the noisiness of the standard error estimate on type 1 error inflation, we replaced the jackknife estimate of the standard error in the denominator of the z-score with the true standard deviation of 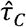 over simulations. This had the effect of increasing significance for simulations for which the standard error was overestimated and decreasing significance for simulations for which the standard error was underestimated. Overall, this reduced type 1 error inflation by a small amount. However, there was still severe inflation in type 1 error even with this correction (Figure 3a,b). Moreover, while in the original analysis, most false positives for the two-sided test were for negative 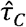 (2492/2591), after correcting the standard error, most false positives were for positive 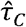 (723/724; Figure S7).

**Figure 3:**
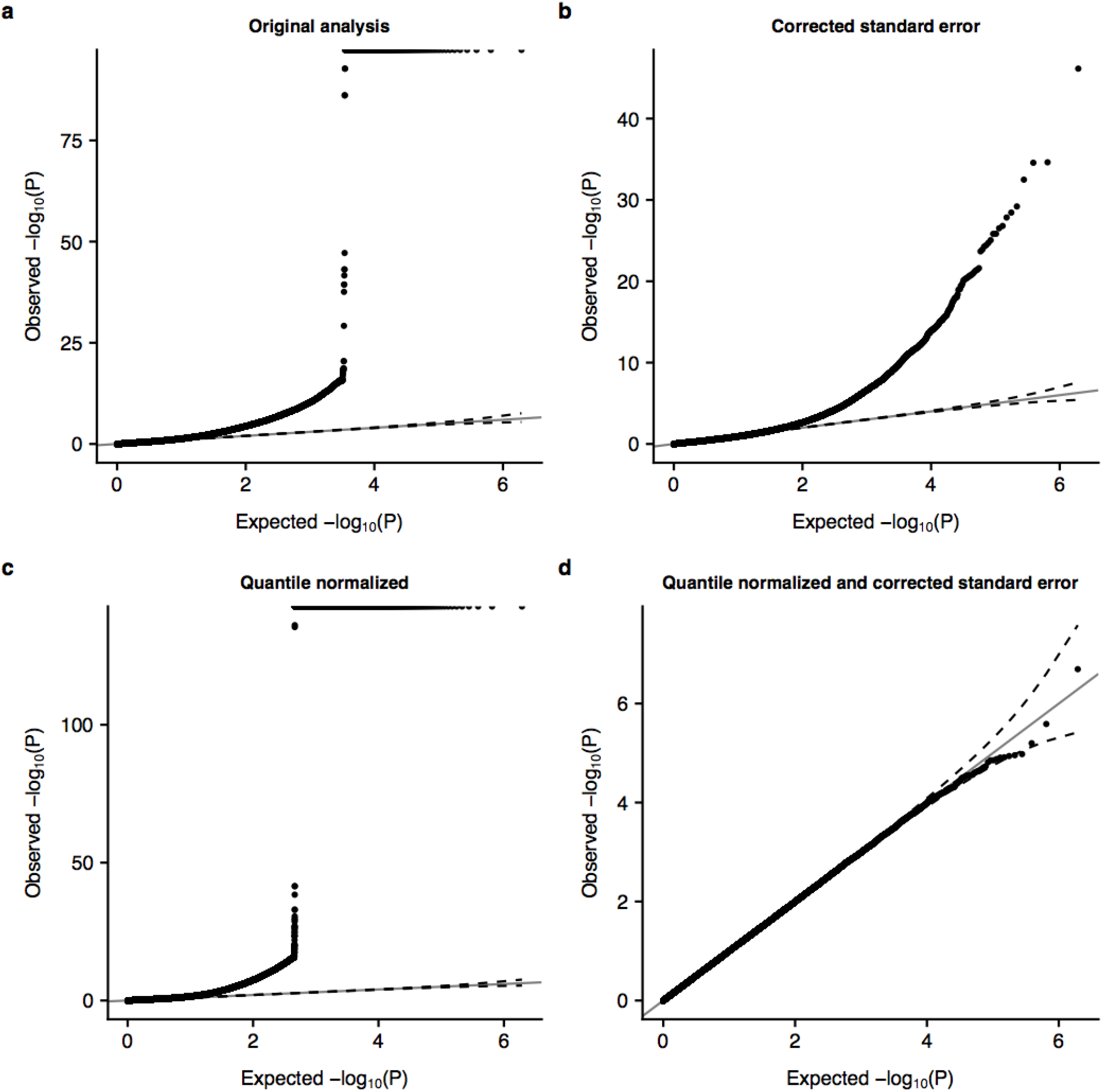
Dependence of type 1 error control on normality of and on the accuracy of the standard error estimate. (**a**) Q-Q plot for the two-sided test of using the jackknife standard error. (**b**) Q-Q plot for the two-sided test of using the corrected standard error. (**c**) Q-Q plot for the two-sided test of the quantile normalized using the jackknife standard error. (**d**) Q-Q plot for the two-sided test of the quantile normalized using the corrected standard error.

Having found that correcting the standard error did not suffice to control type 1 error, we then investigated non-normality of the coefficient estimates as a potential explanation for the type 1 error inflation. To do this, for each annotation, we transformed the estimates of 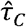 to be normally distributed, preserving the standard deviation. Specifically, for each annotation we first chose 3,750 random samples from a normal distribution with mean zero and standard deviation matching the standard deviation of the 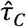 for that annotation; we then quantile transformed the values of 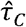 for the annotation to these values. We used the transformed 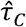 and jackknife standard errors to compute P-values. Overall, this quantile normalization exacerbated the type 1 error (Figure 3a,c, Figure S8a,c). In contrast to the standard error correction, most false positives were for negative 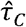both before and after quantile normalization (Figure S9). While neither standard error correction nor quantile normalization sufficed to restore type 1 error control, when applied together they did (Figure 3d, Figure S8d).

We were surprised to find that correcting the standard error left severe inflation of type 1 error and that quantile normalizing 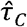 in fact exacerbated type 1 error, but that performing both corrections simultaneously restored type 1 error control. Investigating this further, we found two related phenomena. First, for many annotations there was a positive correlation across independent simulated phenotypes between 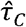 and the standard error estimate, and this correlation tended to be higher for the two least polygenic sets of phenotypes than for the most polygenic set of phenotypes (Figure S10). This correlation resulted in higher standard error estimates and thus more conservative p-values when the coefficient estimate was positive, and lower standard error estimates and thus less conservative p-values when the coefficient estimate was negative. The second phenomenon we observed was that for most annotations that were non-normal (P < 0.05 using a Kolmogorov-Smirnov test for normality), the distribution of 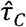 had a right skew (mean > median for 183 out of 191 annotations).

Together, these two phenomena explain several aspects of our results. Because the original distribution of 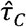tended to be right-skewed, quantile normalizing the 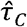 distributions mostly reduced right skew and increased left skew, thus mostly increasing significance for the most significant negative 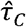. Because of the correlation between 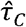 and jackknife standard error estimate, standard error correction had the opposite effect: it mostly increased the significance of positive 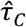 while decreasing the significance of negative 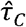. In the original analysis, the dominant contributor to type 1 error inflation was underestimated standard error for negative 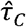. Quantile normalization exacerbated this problem and standard error correction ameliorated it; hence quantile normalization increased type 1 error inflation while standard error correction decreased it. After standard error correction, right-skewed non-normality became the main source of type 1 error, and most false positives were for positive 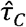.

Finally, we ran MAGMA^9^, a commonly used gene set enrichment method, on our simulated phenotypes and gene-based annotations to determine whether it also resulted in inflated type 1 error. Aggregated across annotations and phenotypes, MAGMA had mildly inflated type 1 error (Figure S11).

## Discussion

We applied a simulation framework to characterize the performance of the block jackknife-based significance testing used in S-LDSC. Using 3750 simulated phenotypes with varying levels of polygenicity, we ran S-LDSC on 94 simulated annotations, the baseline_v1.1 annotations, and 100 real gene sets sampled from MSigDB. For small annotations, we observed significant inflation in the reported one-sided and two-sided p-values for 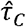 but only mild inflation in the heritability enrichment p-values from S-LDSC. The inflation is due both to the noisiness of the jackknife estimate of the standard error, and to the non-normality of the regression coefficient estimates. This inflation can be remedied by restricting to annotations that overlap at least 1.7% of 0.01 cM blocks for standard analyses (i.e., for the one-sided test of 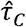 and the two-sided test of heritability enrichment) and 8.3% of 0.01 cM blocks for the two-sided test of 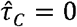. We have implemented a test in the S-LDSC software to warn the user when their annotation does not meet the criteria required to produce statistically valid p-values. For small and/or clustered gene sets, MAGMA may provide better type 1 error control; we leave a thorough investigation of type 1 error for MAGMA to future work.

For small annotations, our simulations showed perfect type 1 error control at P=0.05 but a severe inflation of very small P-values. This highlights the need to assess type 1 error control of new methods by looking not only at a single fixed cutoff, but also by examining the tail. This is particularly important for tools such as S-LDSC that are often used to test a very large number of hypotheses, with stringent cutoffs for significance after multiple testing correction.

It is possible that the true null distribution of regression coefficient estimates for a given annotation could be derived or simulated and then used for hypothesis testing for small annotations. However, our results indicate the null distribution will depend on the genetic architecture of the trait being studied, presenting a challenge. Moreover, derivations will involve higher moments of the genotype matrix, leading to potential difficulties in both computation and reference panel mismatch. Since LD score regression controls type 1 error for annotations that are not very small, we propose here a simple restriction on input instead of a new method for significance testing.

We note three limitations of our work. First, the null simulations performed here are of heritable phenotypes with no enrichment in any functional annotation. We caution that, as noted in earlier work^5,8^, model misspecification such as enrichment in a category not included in the model can also lead to bias and inflated type 1 error. Model misspecification is best addressed by fitting as flexible a model as possible, and will not be fixed by the threshold on 0.01 cM blocks overlapped introduced here. Second, we simulated a range of annotations including both gene sets and sets of random SNPs, and while we believe these annotations represent typical S-LDSC input, we cannot guarantee that our results extend to arbitrary annotations. Third, all annotations tested in this work are binary annotations. We leave characterization of type 1 error for continuous annotations, including those of the baseline-LD model^11^, to future work.

In conclusion, S-LDSC produces well-calibrated p-values when annotations are large and spread throughout the genome, regardless of the level of polygenicity of the trait tested. We recommend performing standard S-LDSC analyses only on annotations that span at least 1.7% of 0.01 cM blocks of the genome, and performing two-sided tests for *τ*_*c*_ = 0 only on annotations that span at least 8.3% of 0.01 cM blocks.

## Data Availability

All data can be found within the manuscript, MSigDB, and UKBiobank.

https://www.gsea-msigdb.org/gsea/msigdb

## Acknowledgements

We thank L. Abbott, S. Gazal, D. Palmer, A. Price, J. Ulirsch, R. Walters, E. Weeks and O. Weissbrod for helpful discussions. HKF is supported by NIH grant DP5 OD024582 and by Eric and Wendy Schmidt. This research was conducted using the UK Biobank Resource.

## Supplemental Figures and Tables

**Table S1:**
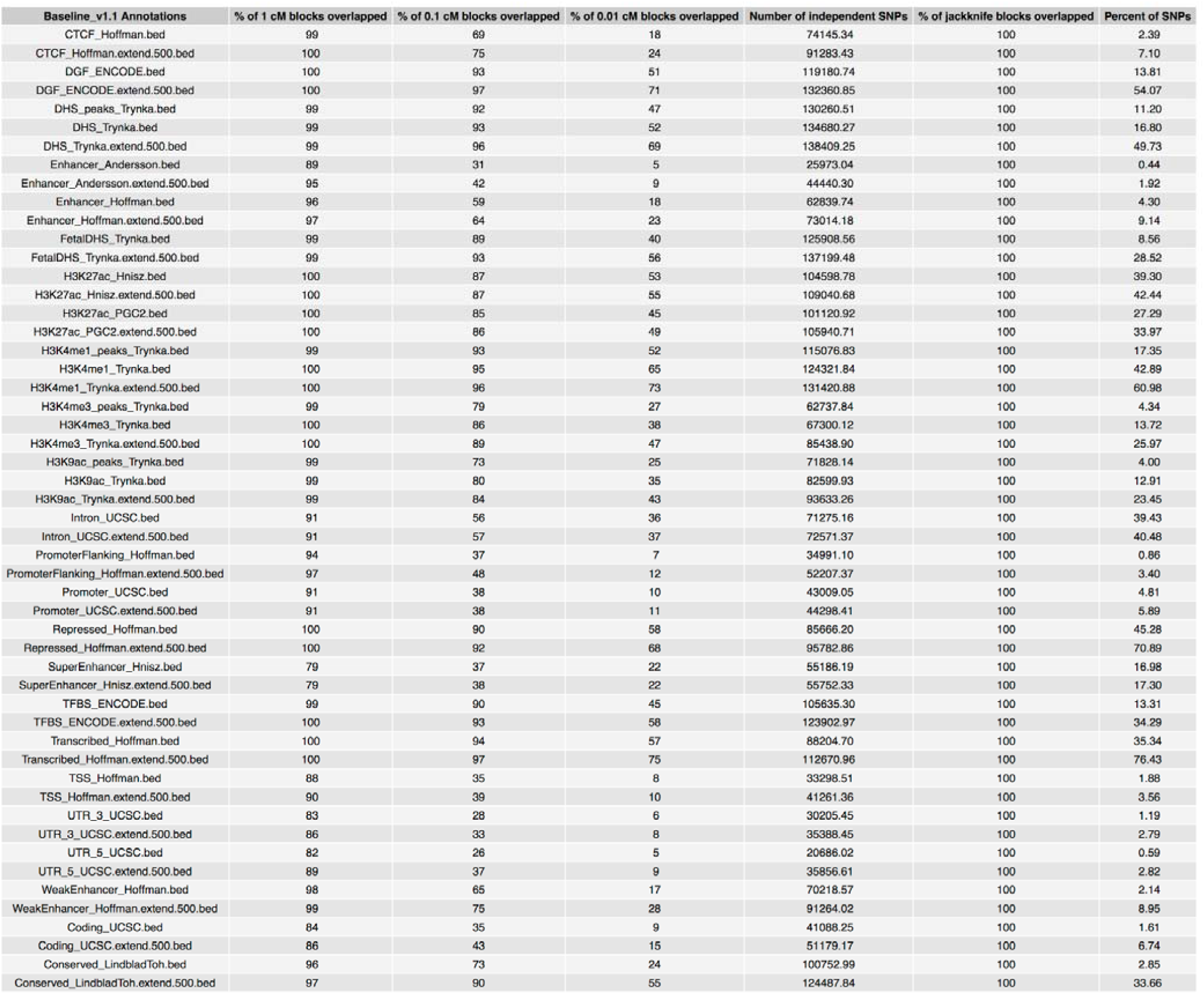
The annotation metrics for the baseline_v1.1 annotations.

**Figure S1:**
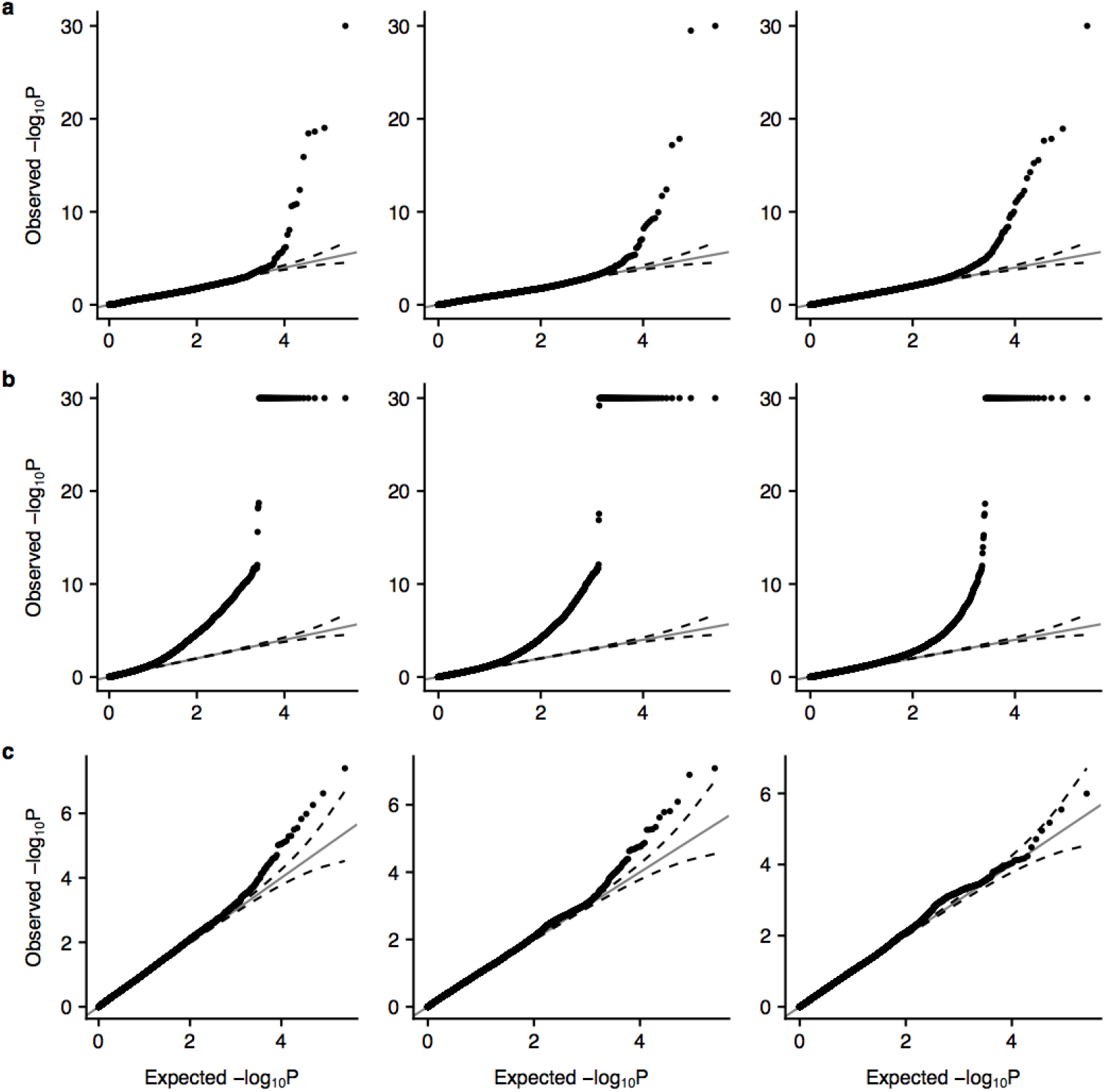
Results are consistent across polygenicities. (**a**) The results from the one-sided test of. (**b**) The results from the two-sided test of. (**c**) The results from the enrichment test. Polygenicity increases from left to right in each case.

**Figure S2:**
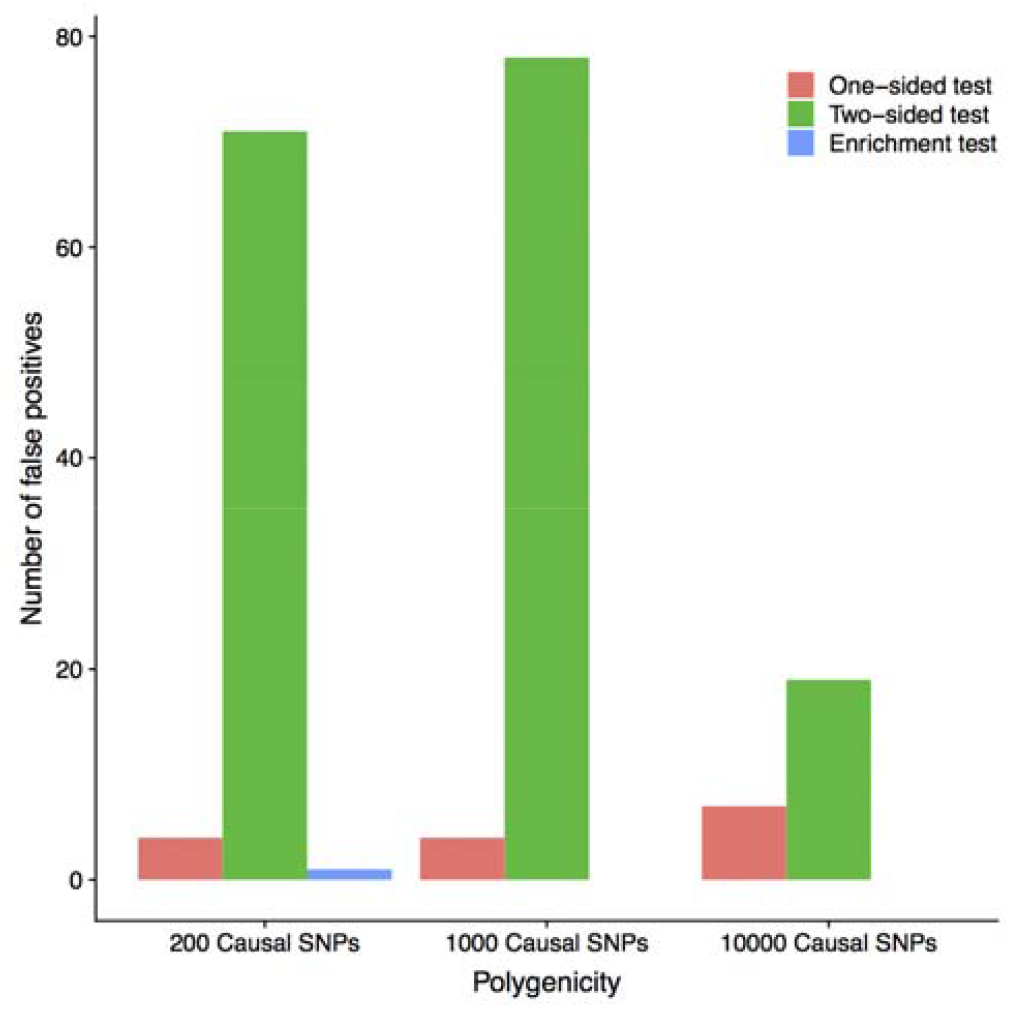
The number of false positives (P<0.05/(3750*246)) for the two-sided test is higher for the two less polygenic sets of phenotypes than for the most polygenic set, while the number of false positives for the one-sided test and enrichment test are stable across polygenicities.

**Figure S3:**
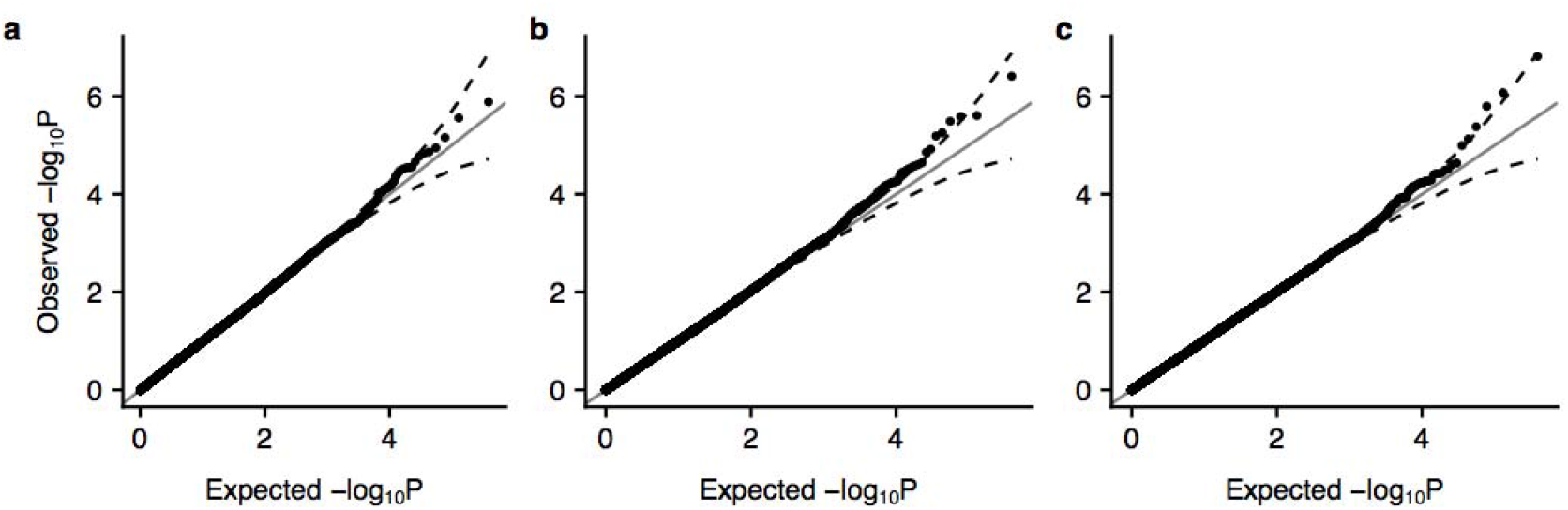
The baseline_v1.1 annotation results have well controlled type 1 error for the (**a**) one-sided, (**b**) two-sided and (**c**) enrichment results.

**Figure S4:**
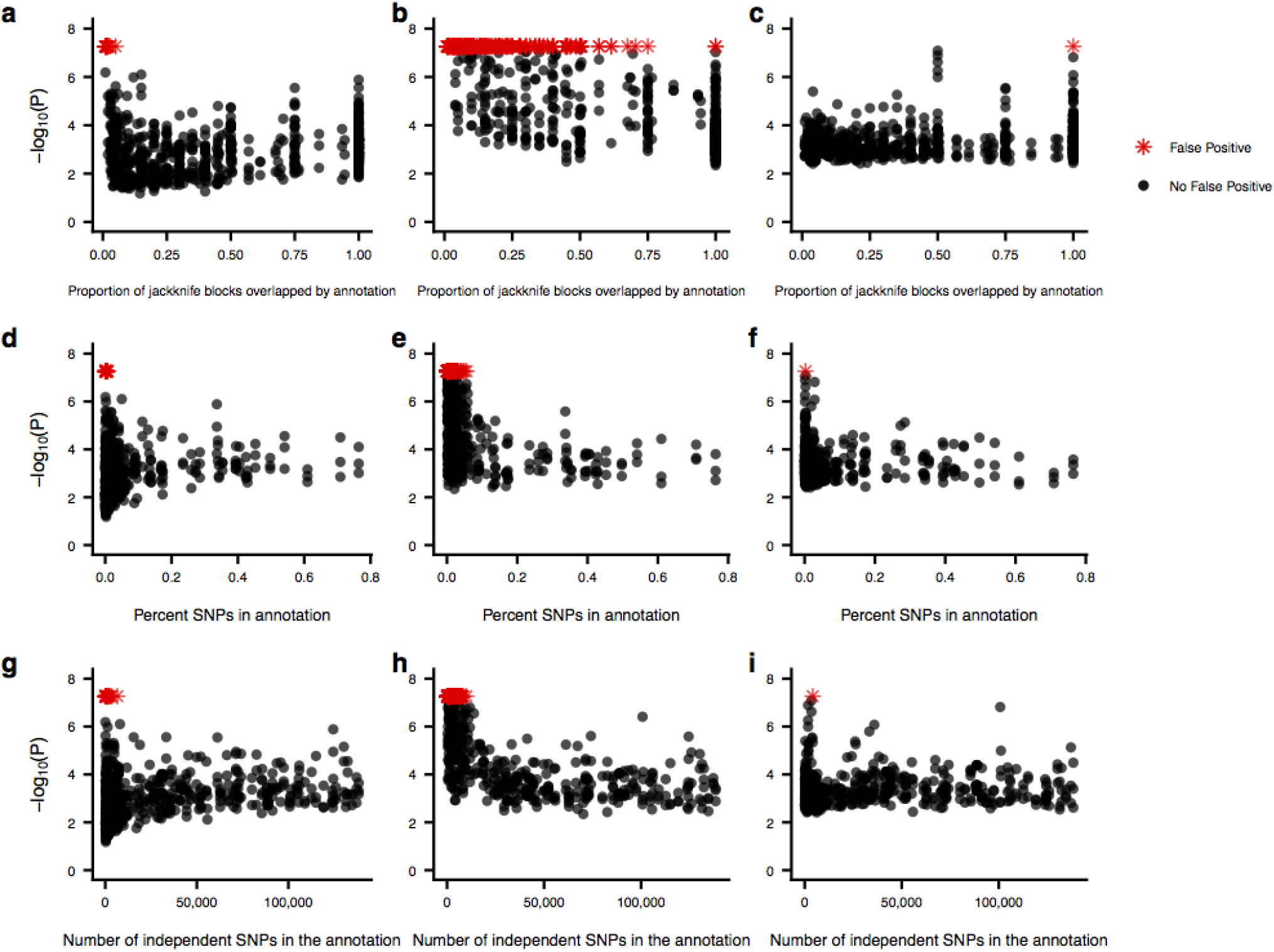
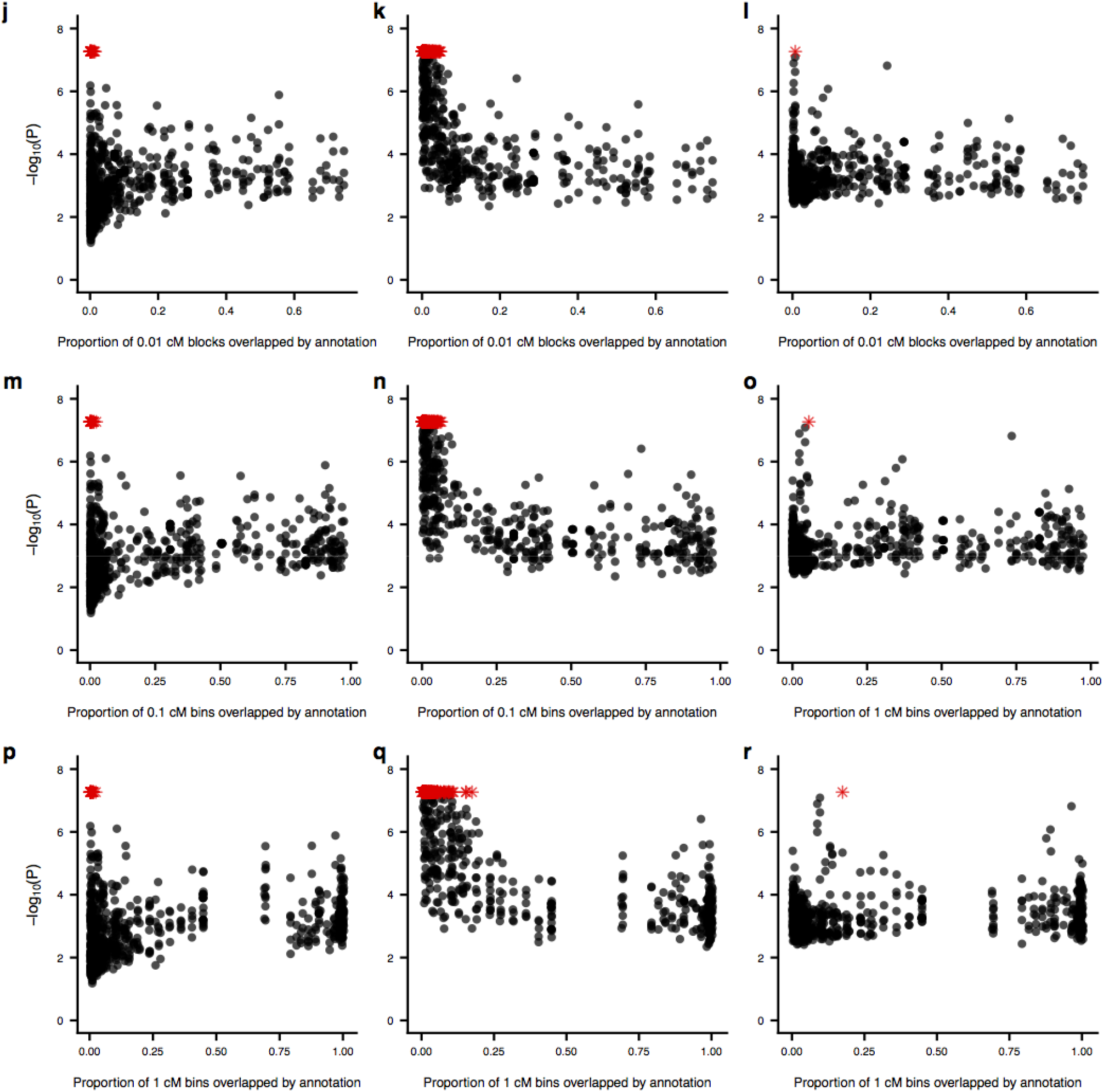
Relationship between significance and the annotation characteristics for the three tests. The annotation characteristics are: (**a-c**) proportion of jackknife blocks overlapped by the annotation, (**d-f**) percent of SNPs in the annotation, (**g-i**) number of independent SNPs in the annotation, (**j-l**) proportion of 0.01 cM bins overlapped by the annotation, (**m-o**) proportion of 0.1 cM bins overlapped by the annotation, (**p-r**) proportion of 1 cM bins overlapped by the annotation. The tests are **(a**,**d**,**g**,**j**,**m**,**p)** one-sided test, **(b**,**e**,**h**,**k**,**n**,**q)** two-sided test, and **(c**,**f**,**i**,**j**,**o**,**r)** test for heritability enrichment. False positives are denoted with red stars.

**Figure S5:**
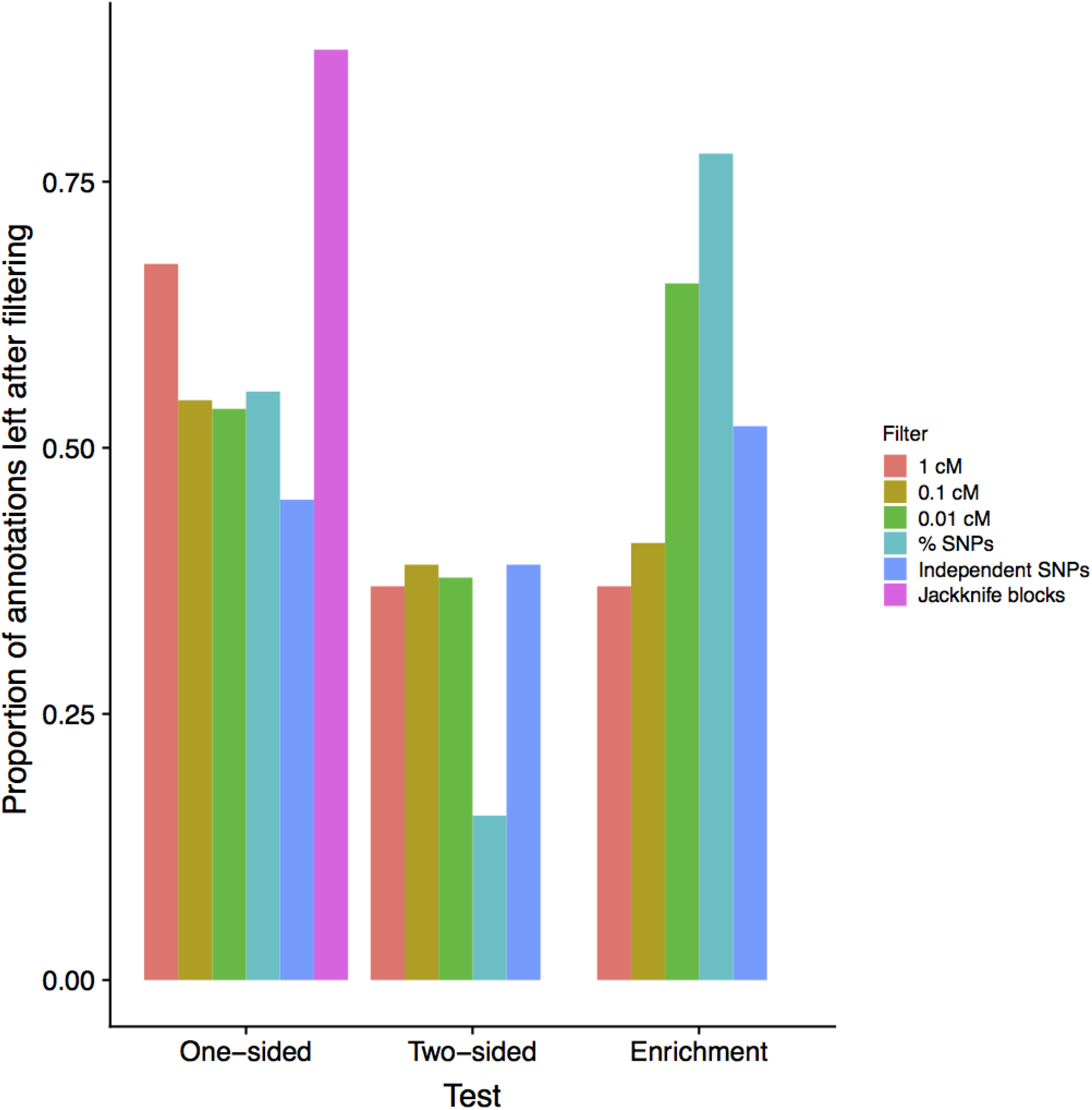
Proportion of annotations remaining after imposing a filter that excludes all false positives. Color denotes the annotation metric used to define the filter.

**Figure S6:**
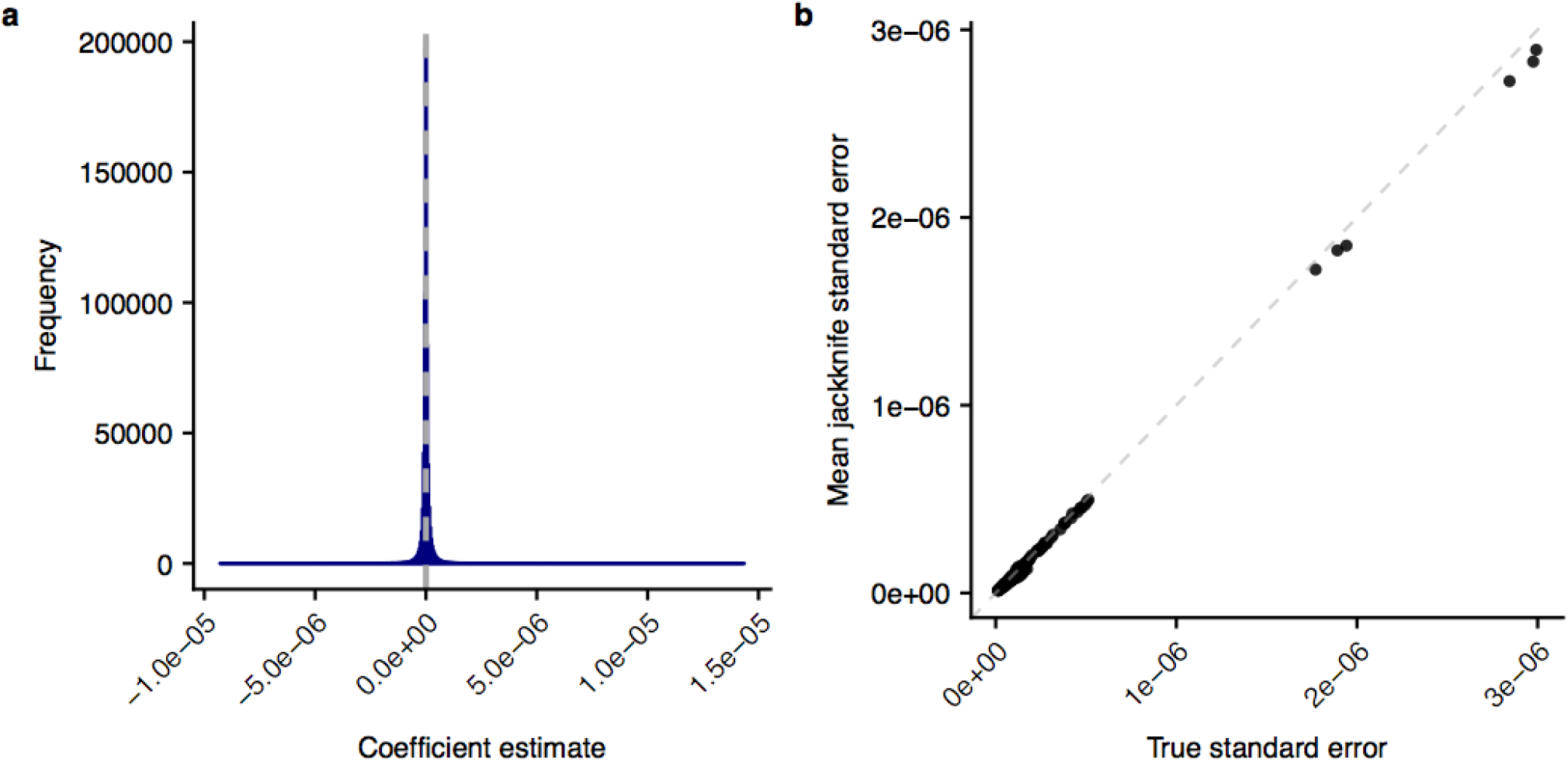
The coefficient estimates and standard error estimates are approximately unbiased. (**a**) Histogram of coefficient estimates over all annotations and phenotypes. The true value for all annotations and phenotypes is =0. (**b**) The true standard error vs. the jackknife standard error for each of the annotations.

**Figure S10:**
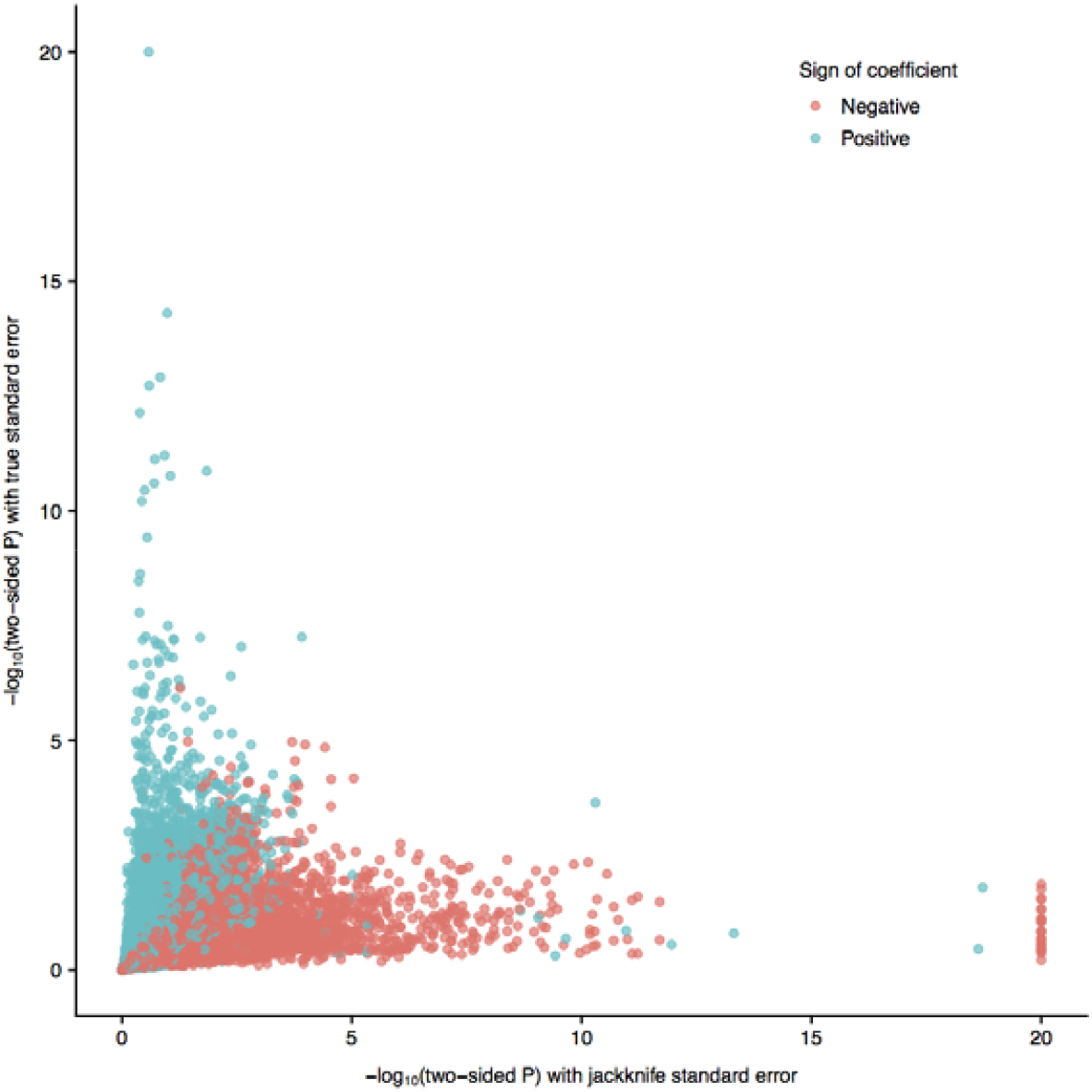
Correcting the jackknife standard error to the true standard deviation tended to increase significance for positive while decreasing significance for negative.

**Figure S8:**
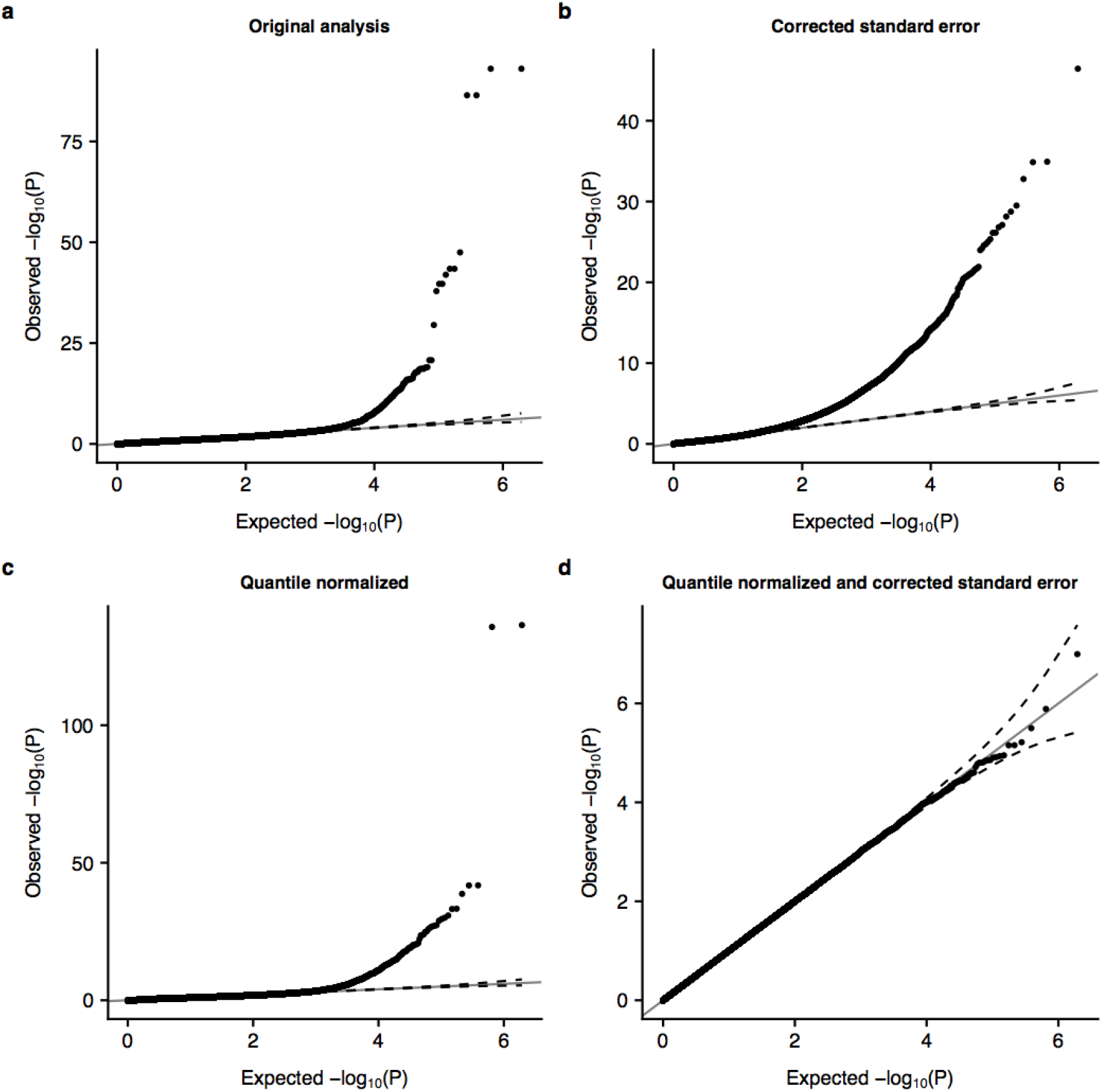
Dependence of type 1 error control on normality of and on the accuracy of the standard error estimate. (**a**) Q-Q plot for the one-sided test of using the jackknife standard error. (**b**) Q-Q plot for the one-sided test of using the corrected standard error. (**c**) Q-Q plot for the one-sided test of the quantile normalized using the jackknife standard error. (**d**) Q-Q plot for the one-sided test of the quantile normalized using the corrected standard error.

**Figure S9:**
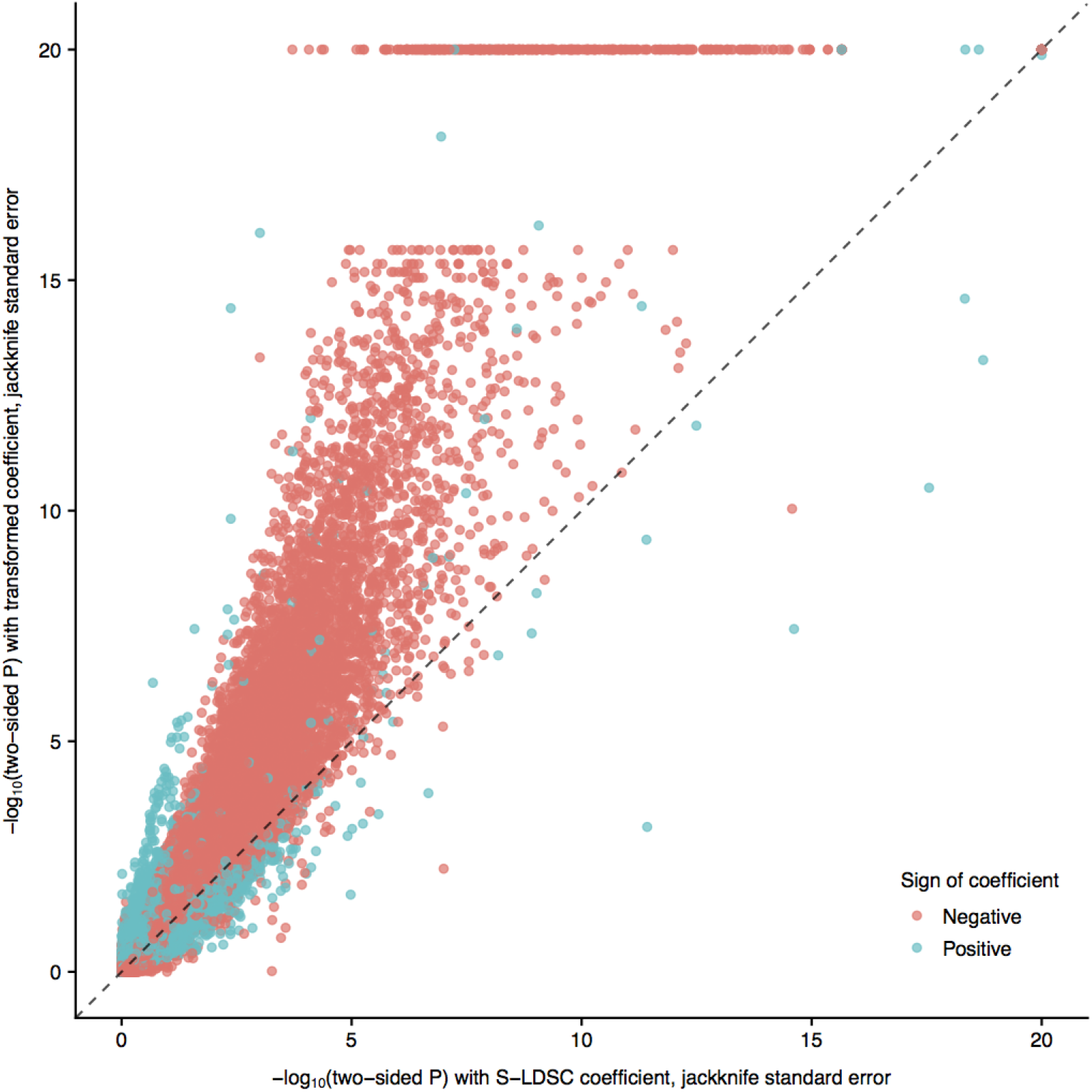
Quantile normalizing the distributions had the effect of mostly reducing right skew and increasing left skew, thus mostly increasing significance for the most significant negative.

**Figure S10:**
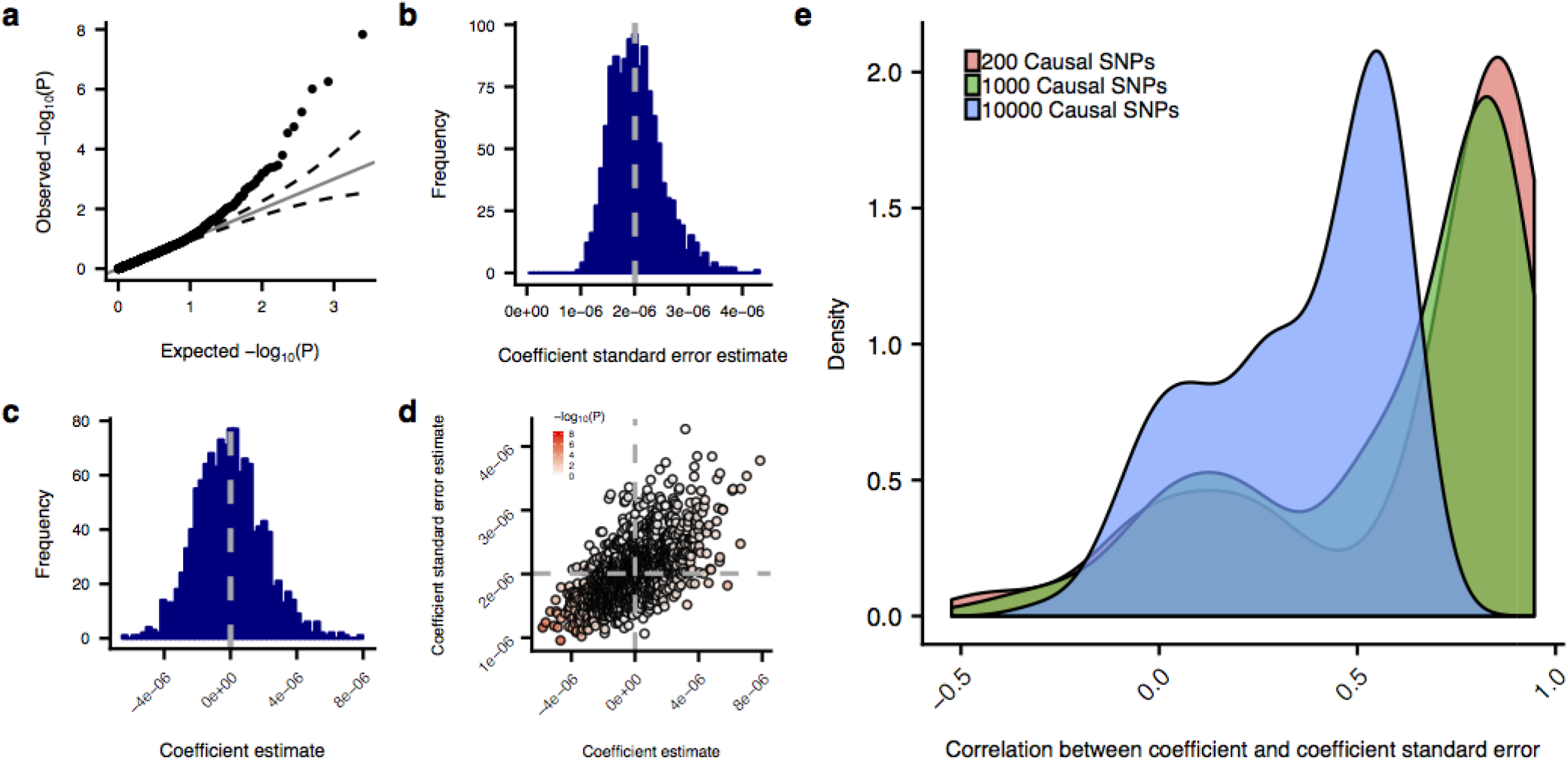
Correlation between and standard error estimate. **(a-d)** Example of inflated type I error in the two-sided test of for an annotation with 0.25% of SNPs overlapping 100 blocks driven by a strong positive correlation between the coefficient estimate and standard error estimate over the mid-polygenic set of phenotypes. (**a**) QQ-plot of p-values from two-sided results. (**b**) Histogram of standard error estimates for over 1250 identical simulations. (**c**) Histogram of over 1250 identical simulations. (**d**) Scatter plot of against the standard error estimate, colored by the -log_10_P of from the two-sided test of. Each point is one of 1250 identical simulations. (**e**) Density of correlations for each annotation between the coefficient and coefficient standard error estimate, colored by the level of polygenicity.

**Figure S11:**
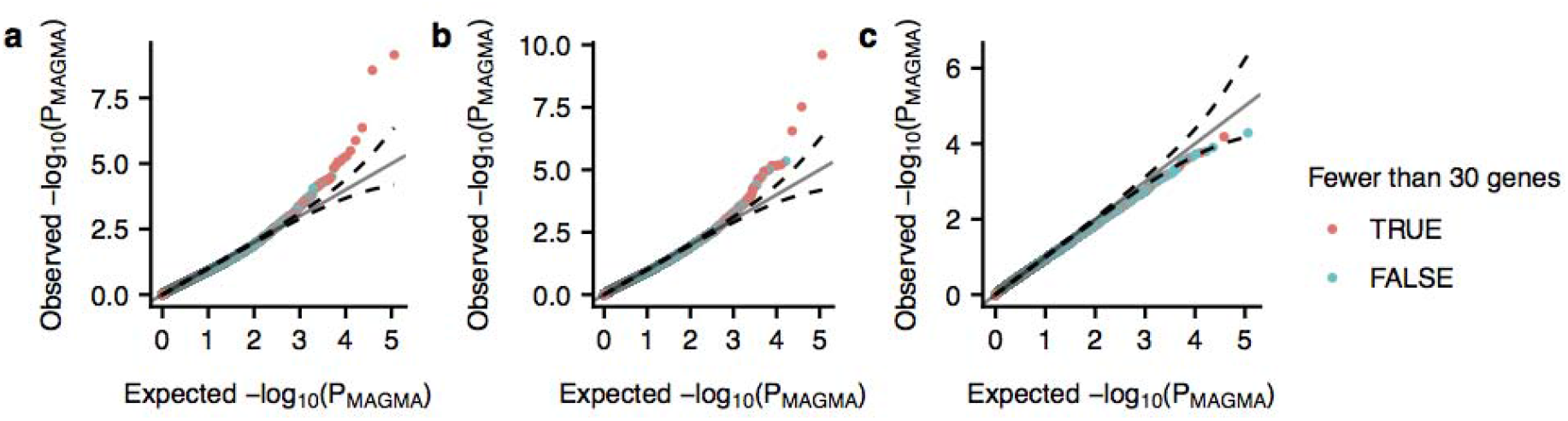
For gene set-based annotations, MAGMA exhibits only mild enrichment. Q-Q plots of MAGMA results on all gene set-based annotations at different levels of polygenicity, ranging from least polygenic to most polygenic (**a-c**). The inflation in panels (a) and (b) appear to be driven by annotations with fewer than 30 genes.

**Table S2:**
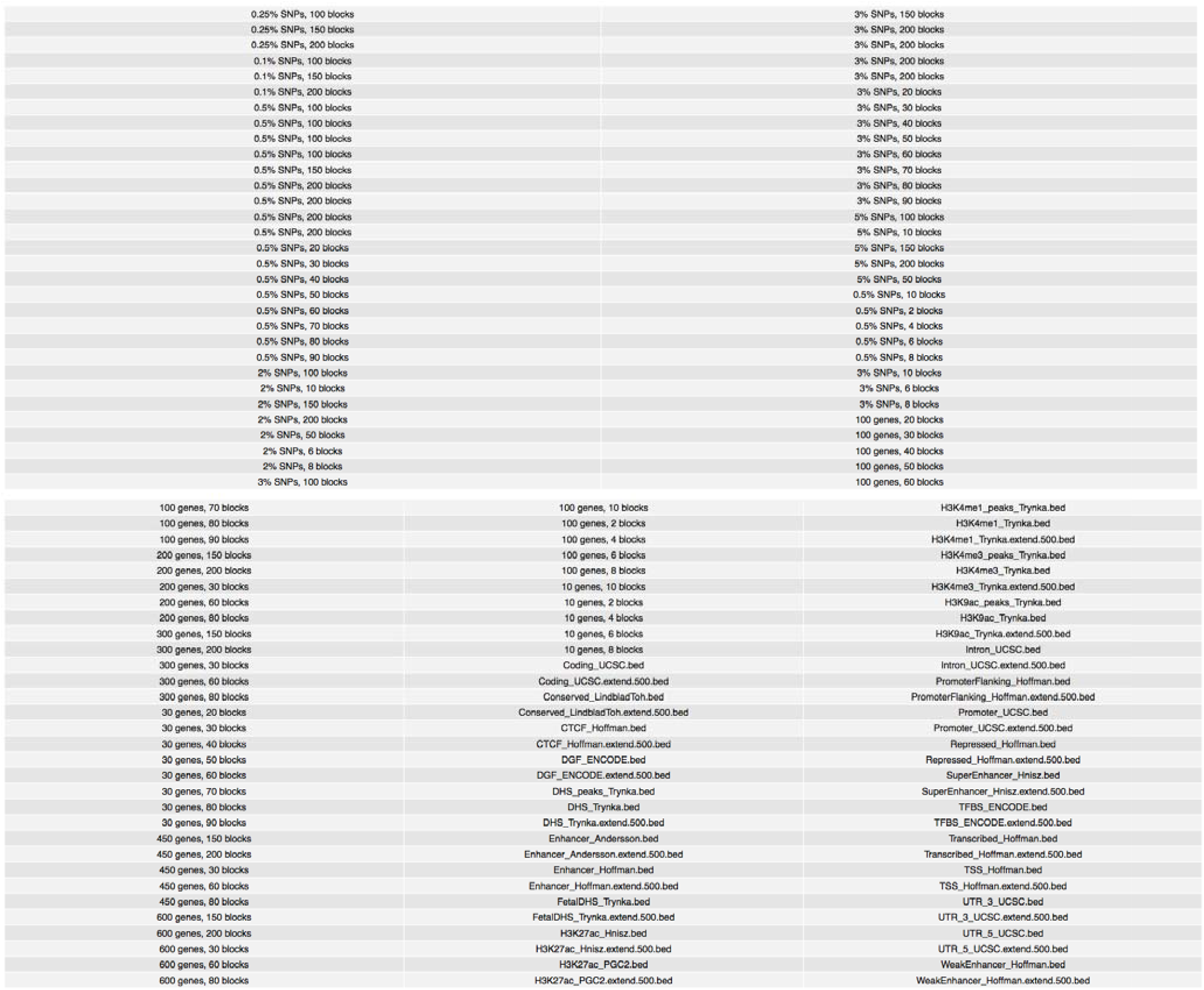
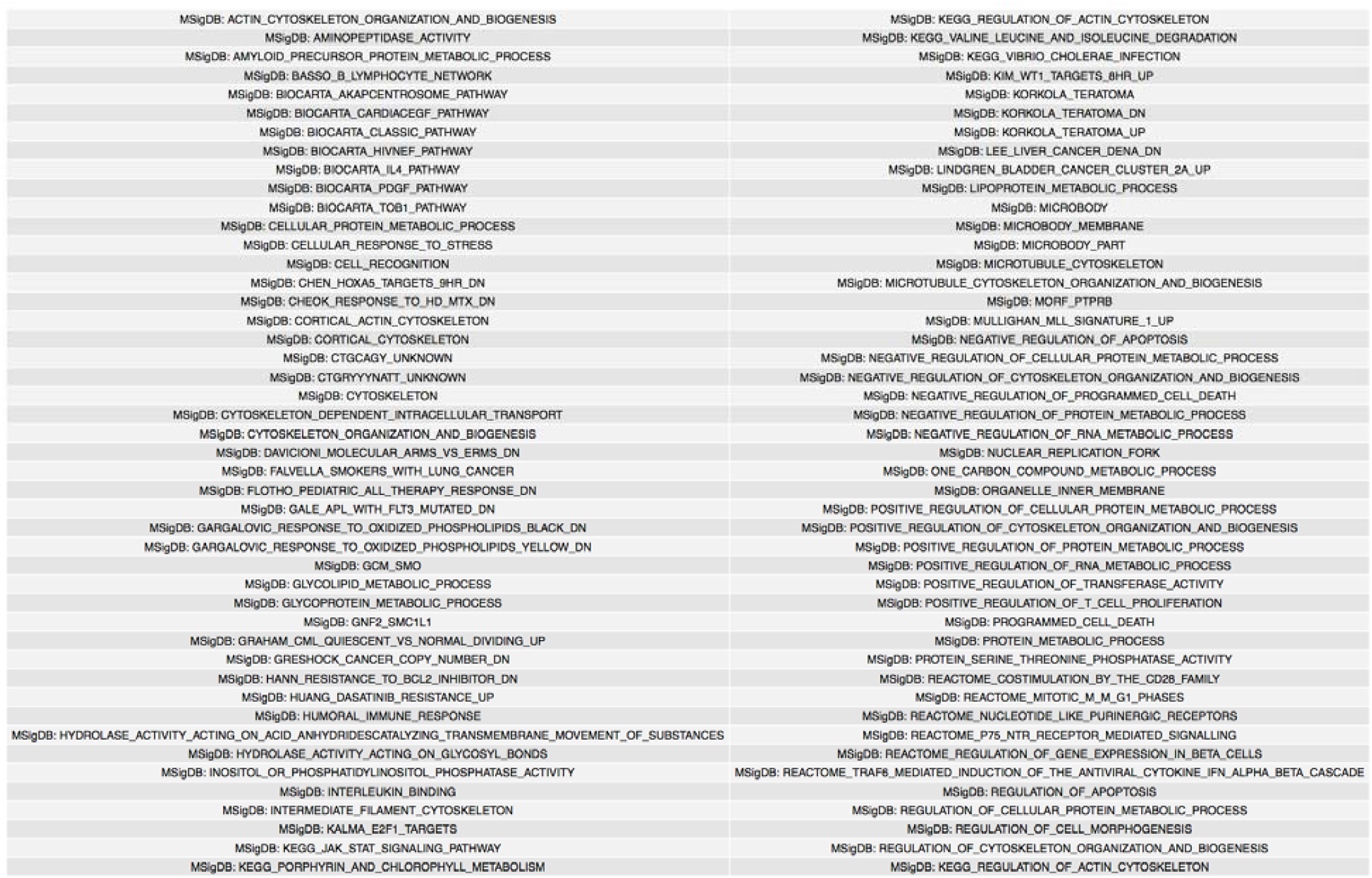
List of 246 annotations used in simulations.

## Notes

### Competing Interest Statement

The authors have declared no competing interest.

### Author Declarations

This research was conducted using the UK Biobank resource, under approved application 31063. All other data analyzed was downloaded from public sources. No original data was collected for this research.

